# Modulation of subcortical activity along the migraine cycle during cognitive executive load

**DOI:** 10.1101/2024.04.03.24305245

**Authors:** Amparo Ruiz-Tagle, Gina Caetano, Ana Fouto, Inês Esteves, Inês Cabaço, Nuno Da Silva, Pedro Vilela, Pedro Nascimento Alves, Isabel Pavão Martins, Raquel Gil Gouveia, Patrícia Figueiredo

**Affiliations:** Instituto Superior Técnico, Universidade de Lisboa, ISR-Lisboa/LARSyS and Department of Bioengineering, Lisbon, Portugal; Faculty of Health Sciences and Nursing, Catholic University of Portugal, Lisbon, Portugal; Learning Health, Hospital da Luz, Lisbon, Portugal; Serviço de Neurradiologia, Hospital da Luz, Lisbon, Portugal; Centro de Estudos Egas Moniz, Faculty of Medicine, Universidade de Lisboa and Hospital de Santa Maria, CHULN, Lisbon, Portugal; Headache Center, Serviço de Neurologia, Hospital da Luz, Lisbon, Portugal; Center for Interdisciplinary Research in Health, Universidade Católica Portuguesa, Lisbon, Portugal

**Keywords:** Menstrual migraine, fMRI, cognition, n-back, migraine cycle

## Abstract

**Objective:** To analyze cognition and brain activation during an executive task in migraine patients studied in the different phases of the migraine cycle, compared with healthy participants.

**Background:** Cognitive difficulties reported during migraine attacks remain poorly understood, despite evidence that the lateral frontoparietal network undergoes reversible disturbances and decreased activation during attacks. Recent findings in resting state functional magnetic resonance imaging suggest that brain areas involved in this network interact with subcortical regions during spontaneous migraine attacks.

**Methods:** Patients with low-frequency episodic menstrual migraine without aura were assessed with functional magnetic resonance imaging while performing a working memory task, along the four phases of natural migraine cycles, including spontaneous attacks, namely: peri-ictal (preictal, ictal, postictal) phases and interictally (in-between attacks). Healthy controls were assessed during the corresponding phases of their menstrual cycles.

**Results:** The protocol was completed by 24 female participants aged 21 to 47 years: 10 with migraine (four sessions each) and 14 controls (two sessions each). Patients and controls showed similar performance on the working memory task and displayed increased brain activity in regions linked to this function, namely the middle frontal gyrus, inferior parietal lobe, and anterior cingulate cortex, during all phases of the migraine/menstrual cycle. Migraine patients exhibited a significant decrease in hypothalamic activity during the postictal phase when compared to perimenstrual controls (*p* = 0.007), interictal (*p* = 0.002) and preictal (*p* = 0.034) migraine phases.

**Conclusion:** Cognitive areas were actively recruited during a working memory task in different phases of the migraine cycle. In addition, migraine patients displayed significantly lower neural activity at the subcortical level in the peri-ictal period. These findings, combined with previous research showing activation in cortical areas, suggest subcortical-cortical interaction during the peri-ictal phases, which may act as a compensatory mechanism when the individual faces a cognitively demanding task during migraine attacks.

## INTRODUCTION

Migraine is the most prevalent brain disorder, affecting at least one in seven people globally^1^, two thirds of whom are women^2^. Although migraine is genetically determined and associated to an excessive brain excitability^3^ it results from an interplay with environmental factors like the hormonal reproductive cycle in women, which explains the disparity in gender prevalence^4^. Imaging studies, namely using functional Magnetic Resonance Imaging (fMRI) have identified abnormal patterns of brain activity occurring during the preictal and ictal phases of migraine attacks^5^. These involve several subcortical structures such as the brainstem, the thalamus, and the hypothalamus, all of which have a pivotal role in initiating and sustaining the attacks^5^.

While headache is commonly acknowledged as the prevailing symptom of migraine, there is a growing recognition that cognitive dysfunction plays a role in all phases (preictal, ictal and postictal) of migraine attacks^6^. An interaction between pain and cognitive processes is in fact expected, given that they share several brain regions including the prefrontal cortex and midcingulate gyrus^7^. A comparative study examining migraine patients and controls during an attentional task, both with and without pain stimulation, observed that only patients exhibited a similar pattern of functional activity, under both conditions, in dorsolateral prefrontal cortex, anterior midcingulate gyrus, and cerebellum, suggesting an interaction between pain and cognition^8^. Only one other previous study from our group analysed patients while performing a cognitively demanding task and found greater activation of prefrontal brain regions involved in pain processing and inhibitory control during spontaneous migraine attacks compared to the interictal state, further supporting the interaction between migraine attacks and cognition^9^.

Other studies of migraine patients using resting-state fMRI found functional connectivity (FC) changes in the Lateral Frontoparietal Network (L-FPN) known to be engaged in cognitive top-down executive functions such as working memory and composed by the Middle Frontal Gyrus (MFG), including the rostral and dorsolateral prefrontal cortex, the Inferior Parietal Lobe (IPL), and the anterior mid-cingulate gyrus (ACG)^9^. For instance, decreased FC of the L-FPN has been observed in a group of patients with episodic migraine during spontaneous attacks when compared to a control group. This detriment of FC correlated inversely with attack frequency (higher FC in L-FPN, lower attack frequency)^10^. Another study that compared interictal migraine participants with controls observed increased FC in L-FPN, which correlated positively with disease duration^11^. Regarding the subcortical structures involved in the pain encoding pathway^12^, the thalamus showed increased FC with the superior parietal lobe during spontaneous attacks when compared to their interictal phase^13^.

In this study we investigate the relationship between working memory performance (using the N-back task) and brain activity (using fMRI) along each phase of migraine cycles including spontaneous attacks. For this purpose, we selected patients with low-frequency episodic menstrual migraine without aura due to their high prevalence (M). Specifically, we aimed to examine attacks associated with menstruation to increase the predictability of spontaneous attacks. We compared this clinical sample to a group of Healthy Controls (HC) in the corresponding stages of the menstrual cycle.

## METHODS

This study is part of a larger research project on brain imaging in migraine (MigN2Treat). It is designed as a prospective, test-retest, within-subject study that included a comprehensive neuroimaging protocol, of which we report here the working memory evaluation using the N-back task-fMRI. The research project also included evaluation of perfusion using arterial spin labelling MRI, microstructure using diffusion MRI, as well as other fMRI protocols, published elsewhere^14–15^. The study was approved by the Hospital da Luz Ethics Committee and all participants provided written informed consent.

### Participants

Women diagnosed with low-frequency episodic menstrual migraine without aura (M), in accordance with the International Headache Society criteria (ICHD 2018)^16^, who had attacks often associated with their menstrual cycle, were recruited during a routine medical appointment at the Headache Outpatient Clinics of Hospital da Luz. HC were women recruited via social media advertisement matching the clinical sample for age and contraception status. Additional inclusion criteria for both patients and controls were: a) age between 18 and 55 years; b) at least 9 years of education; and c) Portuguese as the first language. The exclusion criteria for both patients and controls were: a) diagnosis of a neurologic condition (other than migraine for patients); b) diagnosis of a psychiatric disorder and/or severe anxiety and/or depressive symptoms as indicated by questionnaires; c) daily use of psychoactive medication including migraine prophylaxis for the patient cohort; d) pregnancy, breastfeeding, post menopause, or use of contraception precluding cyclic menses; and e) contraindications for MRI. An additional exclusion criterion for all participants was the evidence of incident brain lesion or structural abnormalities on the structural MRI study.

### Procedure

At recruitment, following informed consent, the neurologist gathered clinical data including disease duration, pain intensity, and attack frequency as well as sociodemographic information. The study protocol then involved four assessment sessions, each corresponding to a distinct phase of the migraine cycle; interictal, preictal, ictal and postictal, as defined by Peng and May 2020^17^, all scheduled in advance based on the menstrual calendar as follows:

- Interictal session: aiming for a date immediately after ovulation. Participants were required to be pain-free for a minimum of 48 hours before the session, and confirmation of the absence of a migraine attack was obtained 72 hours after the session.
- Preictal session: targeting a date near the premenstrual phase. Conducted with a window of 72 hours preceding the onset of a spontaneous migraine attack, with confirmation obtained by contacting patients 72 hours after the session.
- Ictal session: aiming for a date near the beginning of menses. Also, patients were instructed to contact the researchers in the event of a migraine attack outside the perimenstrual period if the pain intensity reached a minimum threshold of 4 points on a 0-10 VAS scale. Ictal sessions occurred at the onset, midpoint, or close to the end of the headache phase, during a spontaneous migraine attack, predominantly during the perimenstrual period. Associated symptoms and clinical details of the ongoing attack were documented. Participants were advised to abstain from using acute anti-migraine medication up to 6 hours before, and during the assessment and scanning process.
- Postictal session: targeting a date shortly after menses, postictal sessions were arranged following a migraine attack whenever the scanner was available, occurring within 48 hours after pain relief from the spontaneous migraine attack.

The first sessions were counterbalanced between peri-ictal and interictal phases across patients.

The study protocol for HC comprised two assessment sessions, chosen at two moments of the menstrual cycle to match the patient sessions, i.e.: one around the menses (perimenstrual) to match the peri-ictal sessions; and another one in the post-ovulation stage to match the interictal session. Also, for HC, the first sessions were counterbalanced between the two phases of the menstrual cycle.

### Assessment sessions

Self-rating questionnaires were administered before scanning, specifically during interictal sessions for M or post-ovulation sessions for HC. Anxiety symptoms were assessed using the Trait (chronic) version of State-Trait Anxiety Inventory (STAI-T)^18^ and depressive symptoms with the Zung Self-Rating Depression Scale (ZSDS)^19^. The impact of migraine was assessed using the Headache Impact Test (HIT-6)^20^ and the Headache Under Response to Treatment (HURT)^21^. Additionally, cognitive symptoms were measured with the Mis-Scog^22^, a nine-item instrument that evaluates subjective cognitive symptoms during migraine attacks, encompassing executive functions and language domains.

### Image acquisition

MRI data were obtained on a 3 Tesla Siemens Vida scanner with a 64-channel radiofrequency receiver head coil. In all sessions, a BOLD fMRI acquisition was performed using a gradient echo-planar imaging (GE-EPI) pulse sequence (TR/TE = 1260/30 ms., simultaneous multislice factor (SMS) 3, GRAPPA acceleration factor 2, voxel size = 2 × 2 × 2 mm³, number of slices = 22, number of volumes = 274).

Structural images were collected on the first session for all participants. We used a T1 weighted sequence (Magnetization Prepared Rapid Acquisition Gradient-Echo, MPRAGE: TR/TE = 2250/2.26 ms., voxel size 1 x 1 x 1 mm³, number of slices = 160).

### Working memory task (verbal N-back)

The verbal N-back task with 0-back and 2-back conditions was used to assess working memory and associated brain activity^23^, using a block design paradigm (Figure 1). The paradigm was built with the Presentation software (Neurobehavioral Systems, USA) and displayed using Nordic NeuroLab hardware, goggles, and response button (www.nordicneurolab.com). The 2-back condition was used as a representative of working memory. Each block had a duration of 24 seconds, with the presentation of a sequence of 12 letters in a pseudorandomized order, for 500 ms. each letter, with a jittered inter-stimulus interval of 1250–1750 ms. (*Average*=1500 ms.). Four blocks of each condition were presented in a fixed alternated order, starting with the 0-back condition. The task had a total duration of 334 seconds (5 minutes and 34 seconds).

**Figure 1.**
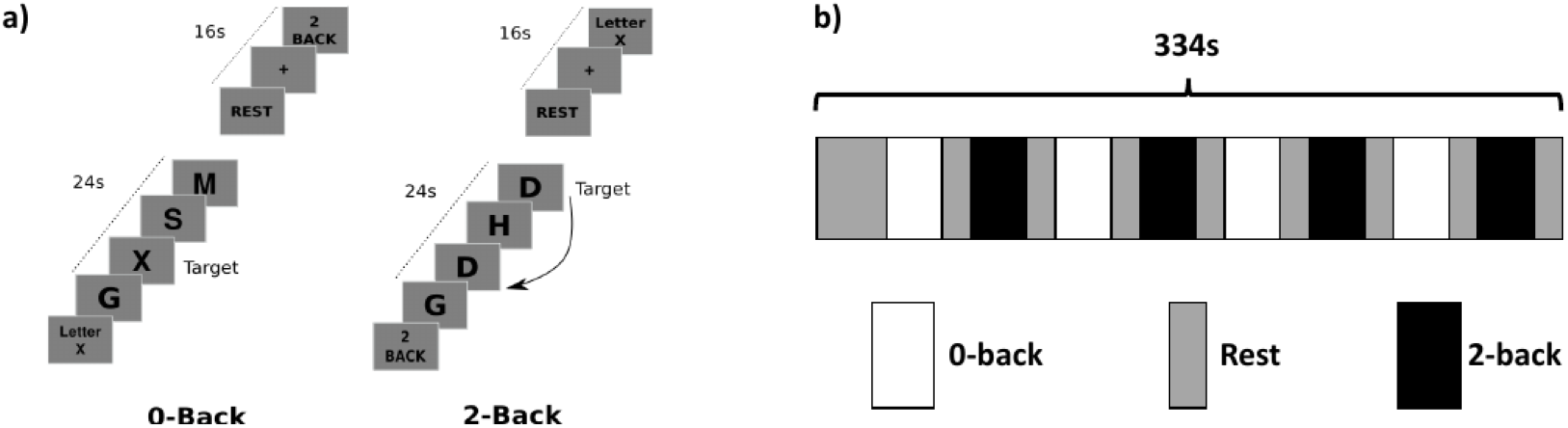
N-back fMRI task used for working memory assessment and associated brain activation. a) Conditions of the N-back task: in the 0-back, the target was every appearance of a randomized pre-selected letter; in the 2-back condition, the target was defined as any letter that matched the one displayed two trials prior in the sequence. b) Schematic of the fMRI N-back paradigm. The task had a total duration of 334 seconds, with four blocks of each condition presented in a fixed alternated order, starting with the 0-back condition. Blocks of both conditions had a duration of 24 seconds with in-between breaks of 16 seconds.

Before entering the scanner, all participants completed a practice trial of the task until they achieved 75 percent accuracy on the 2-back condition. Every session included practice trials. Behavioural measures were also recorded, including Reaction Time (RT), percentage of correct and incorrect responses (when a distractor letter was pressed instead of a target letter).

### Data analysis

The fMRI data was analysed with FSL, the FMRIB Software Library (https://fsl.fmrib.ox.ac.uk/fsl/fslwiki/).

### Pre-processing

Pre-processing included motion correction, fieldmap distortion correction, high pass temporal filtering (frequency cut-off = 100 s.) and spatial smoothing (Gaussian kernel with FWHM = 5 mm). Functional images were registered into the high-resolution anatomical images of the same participant, which were in turn registered to Montreal Neurological Institute (MNI) standard space using nonlinear registration.

### First-level analysis

First-level analysis was performed using a general linear model (GLM) where two sets of explanatory variables (EVs) were generated based on boxcar functions, which corresponded to the visual presentation of the 0-back and 2-back stimulus blocks, respectively, each convolved with the canonical double gamma haemodynamic response function^24^. To tackle motion artifacts, motion outliers were identified and included as confound regressors in the GLM. Additionally, 24 extended head motion parameters were also included as confound regressors in the GLM.

To obtain working memory-related brain activation maps, the contrast 2-back>0-back was computed by subtracting the respective GLM parameter estimates.

### Whole-brain voxel-wise analysis

Group analysis of this contrast was performed voxel-wise for the whole brain, using the FSL tool FEAT/FLAME1. To compare between groups (interictal M phase vs post-ovulation HC and peri-ictal M vs perimenstrual HC) a Two-Sample Unpaired T-test was performed for each comparison. To compare between migraine phases we used repeated-measures ANOVA with four levels (phases) and 12 contrasts for the pairwise comparisons between phases^24^. Statistic images were thresholded using clusters determined by z > 3.1 and a corrected cluster significance threshold of *p* < 0.05^24–25^.

### Region of interest analysis

Region of Interest (ROI) analysis was performed for the following brain areas involved in cognition and migraine pathophysiology, extracted from the Harvard Oxford Cortical Structural Cortical and Subcortical Atlas: bilateral ACC, MFG, IPL, thalamus, and brainstem. Additionally, a ROI was defined for the hypothalamus using the template provided by Pauli and colleagues^26^. A mask was created for each ROI by intersecting the respective anatomical brain areas with the group activation map of the working memory contrast (2-back>0-back), estimated from all sessions included (n = 68). The mean Percentage of BOLD Signal Change (PSC) for the working memory contrast (2- back > 0-back) was computed across each ROI for each participant and session using FSL’s function Featquery.

### Statistical analysis

Non-parametric tests were used for statistical analysis of the ROI PSC values, using IBM SPSS Statistics 26 version: the multiple independent samples Kruskal-Wallis test was used for the comparison between peri-ictal phases and perimenstrual HC, while the Mann Whitney test for two independent samples was used to compare between interictal and post-ovulation groups. To compare between migraine phases the multiple related-samples Friedman’s Two-way analysis of variance by Ranks was used, after normalizing the ROI PSC and behavioral data (correct and incorrect responses percent and reaction times RT). This was done by subtracting the control group mean to the comparative migraine group, i.e., preictal, ictal and postictal – perimenstrual; interictal – post-ovulation. Finally, we conducted Spearman correlations between the normalized 2-back behavioral measures and the ROI PSC values where significant findings were made. Statistical significance was considered at *p* < 0.05 and corrected for multiple comparisons using Bonferroni.

## RESULTS

### 1) Study population and sessions

Thirty-three women were included in the study, of which 18 migraine patients (M) and 16 healthy controls (HC) were matched for age, education, and contraception, and a total of 24 participants having completed the protocol (10 M and 14 HC). Two participants were excluded for incidental MRI findings detected on the first session (one M and one HC). Four sessions were excluded; one lacked task-fMRI recording due to a technical issue, and the other three sessions were scheduled to be preictal, but no migraine attack occurred within 72 hours after the session. Six M participants abandoned the study at different stages. Ten M participants, each contributing four sessions (40 datasets), and 14 HC participants, each with two sessions (28 datasets), completed the study protocol yielding a total of n=68 datasets included in the analysis. The flow chart of the study showing the inclusion of participants in each group and phase is presented in Figure 2.

**Figure 2.**
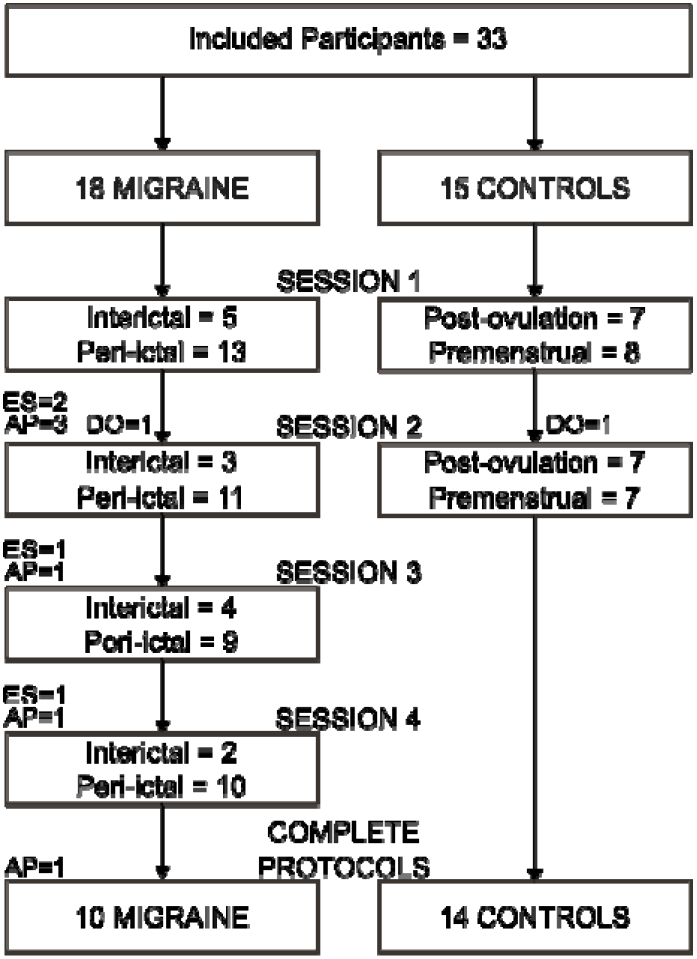
Flow chart of the study. ES: Excluded Session: scheduled preictal with no subsequent migraine attack (n=3), or images not recorded (n=1); AP: Abandoned Protocol (n=6); DO: Drop Out due to incidental MRI findings (n=2).

The final M and HC participants were comparable in terms of age, education, anxiety, and depression scores (Table 1). Both participant groups fell in the category of “Moderate anxiety” (38-44 points) according to the STAI-T and reported “No depression” scores 20-39 in the ZSDS. Out of the 10 M analyzed, four were taking contraception pills and two had the copper Intrauterine Device (IUD), while three HC were under oral contraceptive medication. For the M group, an average score of 23 was obtained in the HIT-6, indicating little to no overall impact of headaches in their lives. The HURT questionnaire yielded a mean score of 8.6, suggesting the existence of burden due to the migraine condition. The Mig-SCog questionnaire had an average score of 9 points with a minimum of 4, indicating perceived cognitive impairment during attacks in all M participants.

**Table 1.** Demographic and clinical data, descriptives and questionnaires scores for both patients (M) and controls (HC). Values are depicted as Median (IQR).

| Parameters | M | HC | p value |
| --- | --- | --- | --- |
| N | 10 | 14 | -- |
| Age (years) | 38.0 (15) | 30.5 (13) | .154 |
| Literacy (years) | 16.5 (1) | 17.0 (1) | .752 |
| STAI-T score | 38.0 (6) | 39.5 (7) | .585 |
| ZDS score | 37.5 (12) | 36.0 (11) | .371 |
| Disease duration (years) | 12.0 (17) | -- | -- |
| Attack frequency (per month) | 2.0 (2.9) | -- | -- |
| Average pain intensity | 7.0 (1.9) | -- | -- |
| HIT-6 score | 21.0 (5) | -- | -- |
| HURT score | 9.0 (2) | -- | -- |
| Mig-Scog score | 8.5 (7) | -- | -- |

Non-parametric test Two Independent-Samples Mann-Whitney with significance set at $p < 0.05$ . STAI-T: State Trait Anxiety Inventory-Trait, ZDS: Zung Depression rating Scale. HIT-6: Headache Impact Test HURT: Headache Under Response to Treatment. Mig-SCog: Migraine Subjective Cognitive impairment.

Table 2 provides a summary of migraine attack associated symptoms and preictal and postictal timings before and after the attacks, respectively. All ictal M patients experienced different levels of photophobia or phonophobia and difficulties in concentration. Some also reported having nausea and sensitivity to movement at the time of examination (Table 2). Ictal sessions were performed either close to the onset, middle or near attack resolution.

**Table 2.** Migraine attack and phases’ description.

| **M phase characterization** | **Median (IQR)** |
| --- | --- |
| **Ictal** |  |
| Time since attack onset (hrs.) | 16 (23.4) |
| Pain intensity* | 6 (4.0) |
| Photophobia / phonophobia* | 3 (4.0) |
| Movement sensitivity* | 5 (6.0) |
| Nausea / sickness* | 1 (3.0) |
| Cognitive difficulties* | 6 (3.0) |
| **Preictal** |  |
| Time before attack onset (hrs.) | 39 (22.3) |
| **Postictal** |  |
| Time after attack resolution (hrs.) | 18 (20.3) |

\*Visual Analogue Scale (VAS) (0-10).

### 2) Working memory (verbal 2-back task) behavioral measures

Behavioral measures from two M sessions (one preictal and one ictal) and four HC sessions (one perimenstrual and 3 post-ovulation) were not recorded because of technical problems with the experimental setup. We analyzed the data corresponding to 62 out of 68 sessions, depicted in Figure 3.

**Figure 3.**
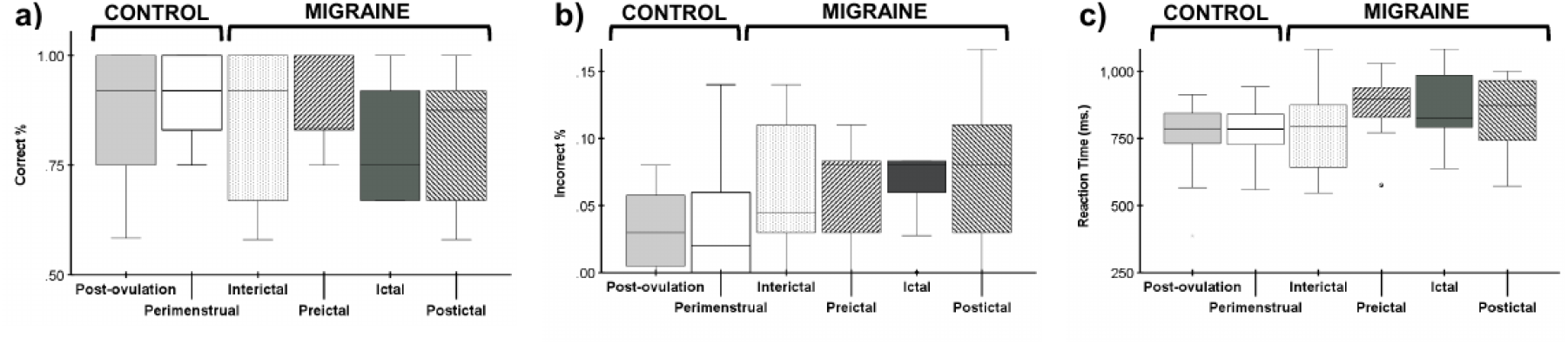
Distribution of behavioral measures across sessions of Migraine and Healthy Controls. a) Percentage of correct responses. b) Percentage of incorrect responses. c) Reaction Time (RT) in milliseconds.

### Migraine and Controls

The performance of all participants in the working memory task was consistent with their age and educational level. No significant differences were found between M interictal and HC post-ovulation for any of the behavioral measures, which includes percentage of correct and incorrect responses, and RT; *p* = 0.771; *p* = 0.409, and *p* = 0.833, respectively. The same was found for the comparisons between M peri-ictal and HC perimenstrual, for percentage of correct (*p* = 0.222), incorrect responses (*p* = 0.054), and RT (*p* = 0.487). HC perimenstrual and post-ovulation also did not yield significant differences in any of the behavioral measures, namely, percentage of correct responses (*p* = 0.395), incorrect responses (*p* = 0.471) and RT (*p* = 0.721).

### Migraine phases

The working memory performance in M sessions declined during peri-ictal phases relative to the interictal phase, yet the decrease was not significant (*p* = 0.820; *p* = 0.199; *p* = 0.348, for the percentage of correct and incorrect responses, and RT, respectively).

### Learning Effect

To enable the comparison of the four phases of the migraine cycle within each M participant, our study design included four repeated measures of the n-back task. Subsequently, an analysis was carried out to ascertain the presence of a learning effect and, if confirmed, to explore its potential influence on our observations. A one-way Analysis of Covariance (ANCOVA) was conducted, with migraine phase as factor and session order included as a covariate. There was no significant interaction observed for the percentage of correct responses (*p* = 0.348) or incorrect responses (*p* = 0.690), nor for RT (*p* = 0.056).

## 3) Working memory (verbal 2-back task) brain activation

### Whole-brain voxel-wise analysis

The group-level brain activation map for the working memory contrast (2-back > 0- back) is presented in Figure 4. This whole brain voxel-wise analysis across all sessions of both patients and controls revealed significant activation during working memory.

**Figure 4.**
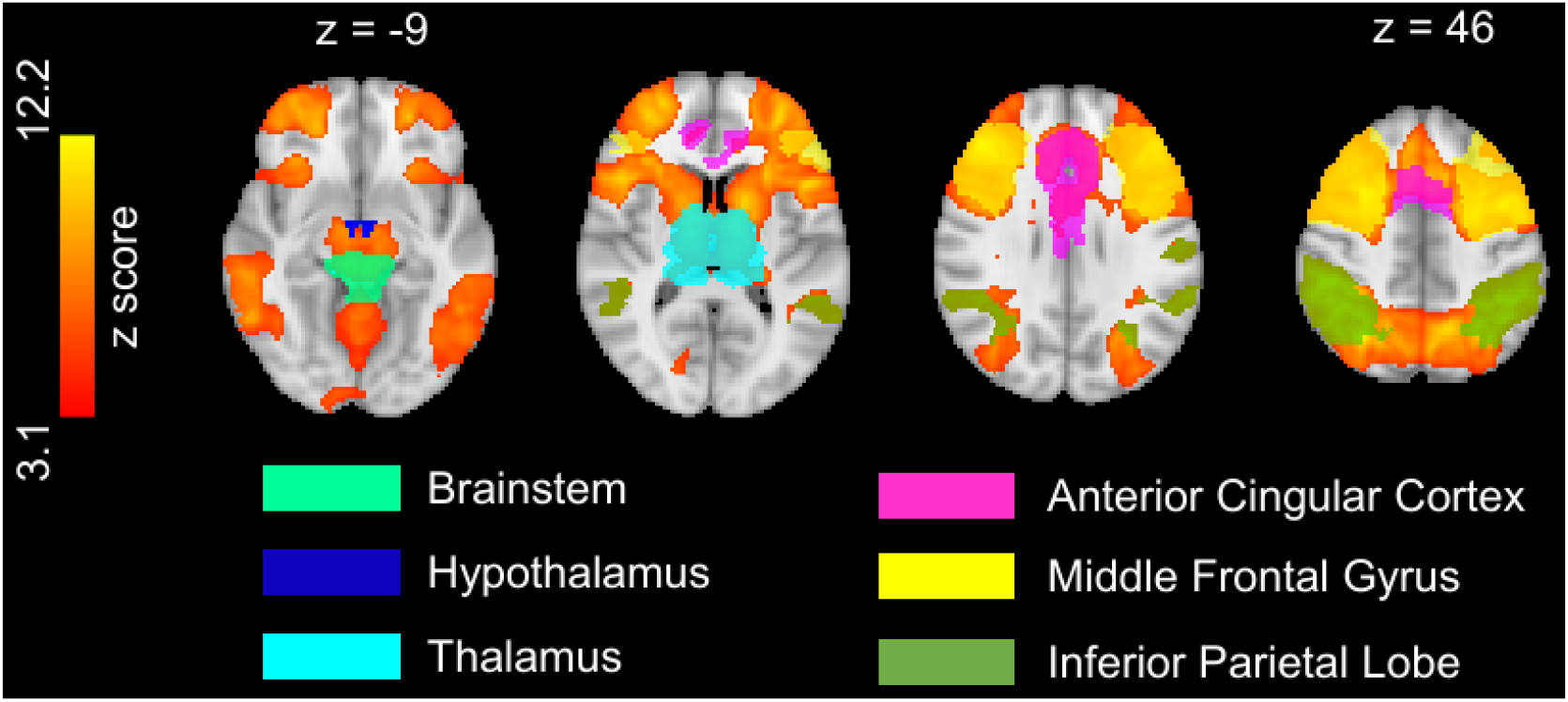
Group level brain activation map for the working memory contrast of the N-back task (2-back > 0-back) (z stat map thresholded with cluster-based correction at z > 3.1, overlaid on the MNI structural template). Representative axial slices are shown on the top from left to right. Areas selected for Region of Interest Analysis are displayed with color legends for the brainstem, thalamus, hypothalamus, ACG, MFG and IPL.

There were no statistically significant differences identified voxel-wise between HC and M. Similarly, voxel-wise comparisons between the different migraine phases did not yield statistically significant differences. HC comparison between menstrual phases was also not significant.

### Region of interest analysis

The ROI analysis revealed a main effect in the hypothalamus when comparing M peri- ictal and HC perimenstrual, with *p* = 0.008, post hoc analysis showed that the postictal drop in BOLD drove this difference with an adjusted *p* of 0.007. Remaining ROIs were not significant for this comparison in ACC *p* = 0.517, MFG *p* = 0.696, IPL *p* = 0.424, thalamus *p* = 0.283 and brainstem *p* = 0.240. No significant differences were found when comparing M interictal and HC post-ovulation, in the ACC (*p* = 0.558), the MFG (*p* = 0.598) and the IPL (*p* = 0.907), brainstem (*p* = 0.219), hypothalamus (*p* = 0.682), or thalamus (*p* = 0.770). Statistical differences were also not observed among HC sessions, perimenstrual and post-ovulation, with ACC *p* = 0.638, MFG *p* = 0.683, IPL *p* = 0.875, thalamus *p* = 0.363, brainstem *p* = 0.778, nor hypothalamus *p* = 0.433. ROI analysis across all subjects and sessions are depicted in Figure 5.

**Figure 5.**
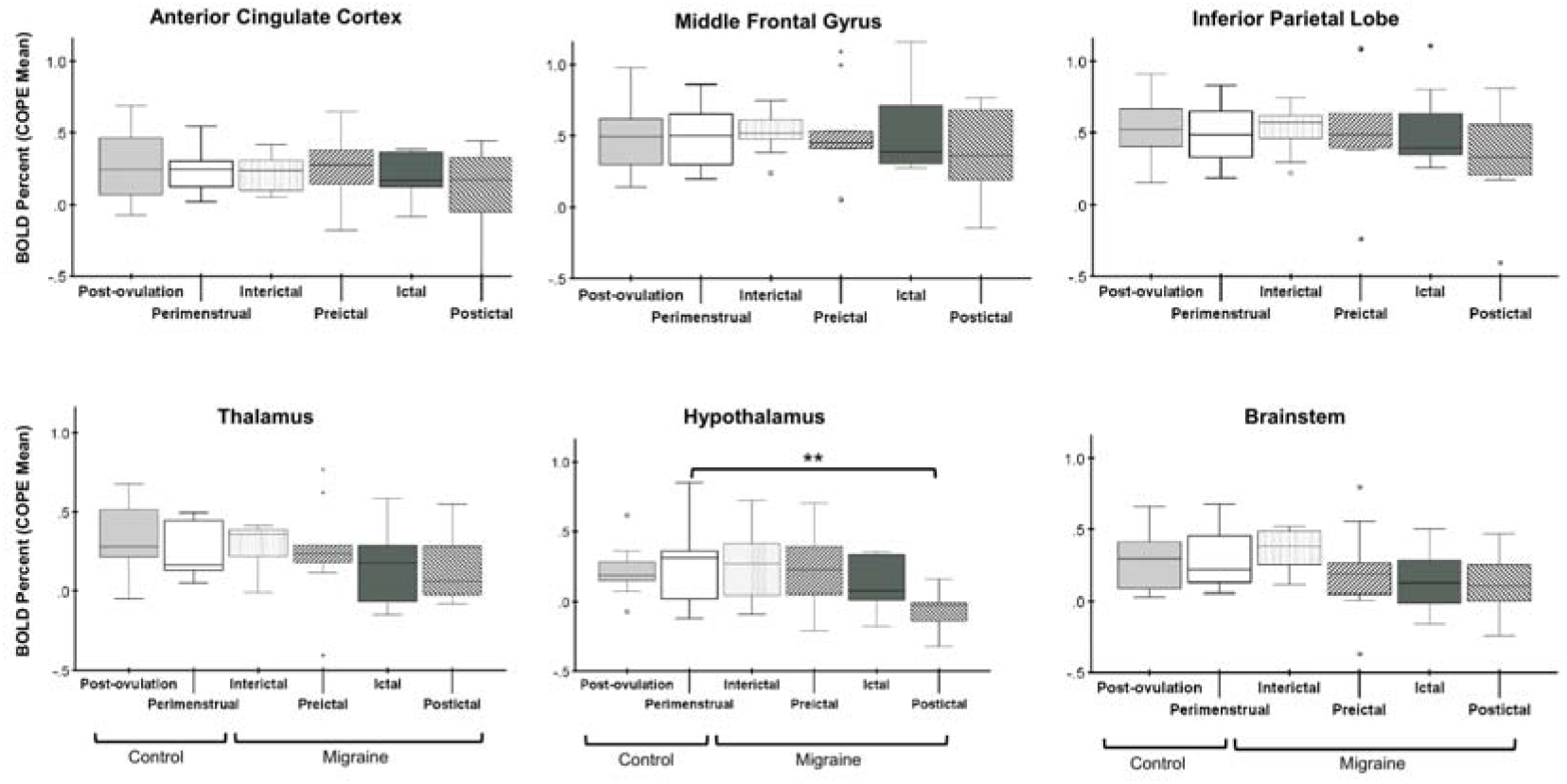
Percent BOLD signal change (COPE mean) values were extracted to compare their distribution between phases. Preictal, ictal and postictal migraine phases were compared to HC perimenstrual (with Kruskal Wallis for multiple independent samples) where only the hypothalamus was significant with *p* = 0.007. No significant differences were found in any ROI in between M interictal and HC post-ovulation (with Mann Whitney for two independent samples). Multiple comparisons were performed using Bonferroni.

Comparisons in-between migraine phases with the normalized PSC values revealed a main effect in the ACC (*p* = 0.015), hypothalamus (*p* = 0.001), brainstem and thalamus (*p* = 0.034 and *p* = 0.045) respectively. Post-hoc analysis revealed that the postictal phase specifically drove this difference; in the hypothalamus when compared to the interictal, (*p* = 0.002) and preictal (p = 0.034), and in the ACC when compared with the preictal (*p* = 0.019). MFG and IPL did not yield statistical differences with *p* = 0.356 and 0.293, respectively. Distribution of normalized values across migraine phases are depicted in Figure 6.

**Figure 6.**
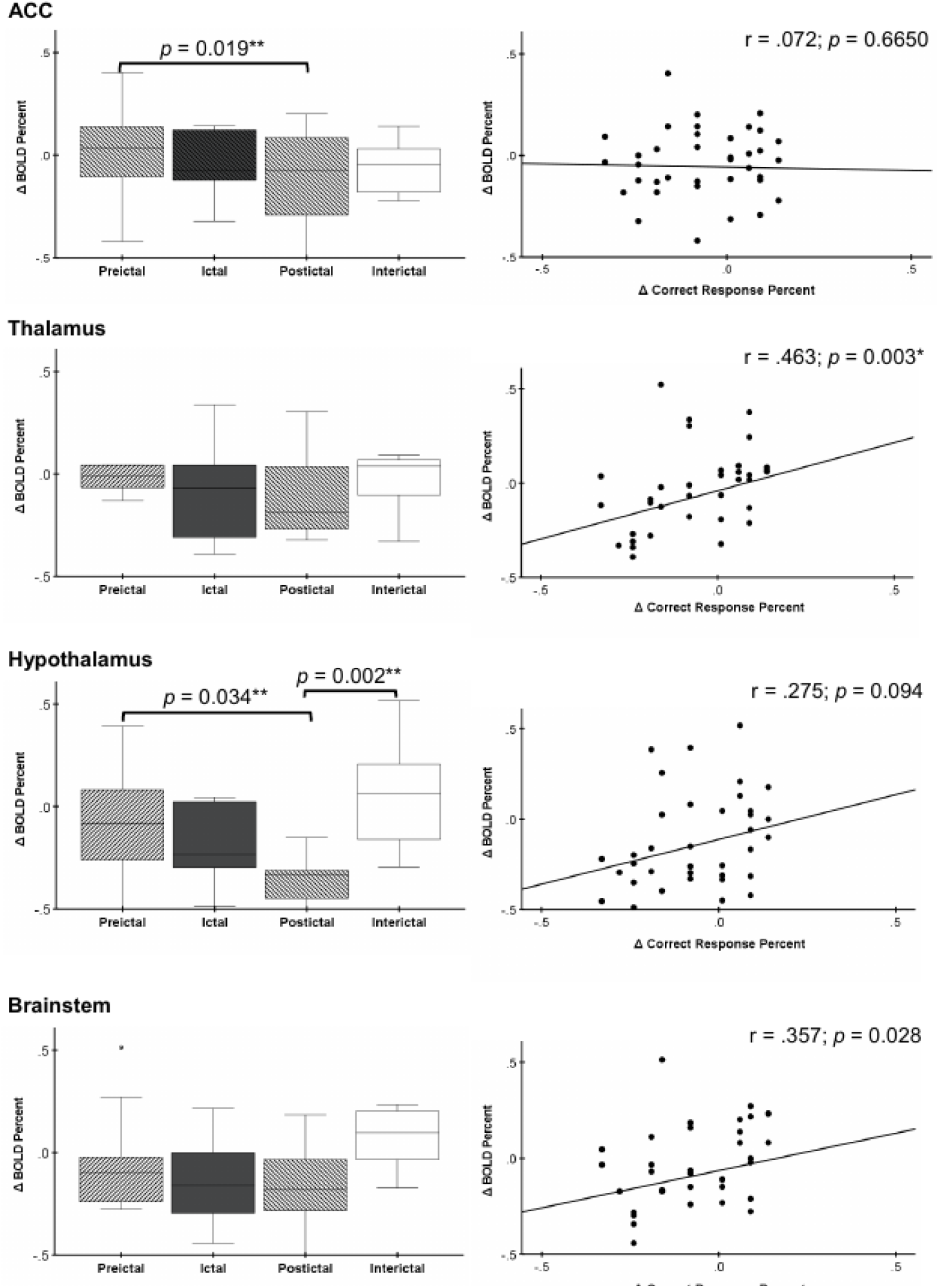
Region of interest analysis with the difference in BOLD percent signal change during working memory (2-back>0-back) in patients relative to controls, during each of the four phases of the migraine cycle, for the Anterior Cingulate Cortex (ACC), thalamus, hypothalamus, and brainstem. Boxplots represent distributions across patients (N=10) and adjusted *p* values refer to the post hoc statistical analysis across phases after main effect with multiple related samples Friedman nonparametric test. The scatter plots of the PSC with the percentage of correct responses in the 2-back condition are shown on the right, indicating the Spearman correlation and the respective *p* value with significance level set at 0.005 after Bonferroni correction for multiple comparisons.

## 4) Correlations

Normalized 2N-back behavioral measures and PSC values of ROIs that yielded statistically significant differences were correlated, namely the ACC, thalamus, hypothalamus and brainstem. Brain activation in all three subcortical structures showed a general positive correspondence with 2-back correct responses percent, and a negative association with incorrect responses percent and RT. Only the correlation between the thalamus and percentage of correct responses was significant after multiple comparisons correction with *r* = .463 and *p* = 0.003. Neither the brainstem and percentage of correct responses (*r* = .357, *p* = 0.028), incorrect responses (*r* = -.321, *p* = 0.049) and RT (*r* = -.207, *p* = 0.212), nor the hypothalamus with percentage of correct responses (*r* = .275; *p* = 0.094), incorrect response (*r* = -.208; *p* = 0.211) and RT (*r* = -.179; *p* = 0.282) were significant. Correlation between the thalamus with percentage of incorrect responses (*r* = -.346; *p* = 0.034) and RT (*r* = -.276; *p* = 0.093) were also not significant. ACC did not yield significant correlation with percentage of correct responses (*r* = .072; *p* = 0.665), incorrect responses (*r* = -.101; *p* = 0.548), nor RT (*r* = .105; *p* = 0.529). Bonferroni correction was performed for 12 correlations and significance level set at *p* < 0.005. Correlations for percentage of correct responses are depicted in Figure 6.

## DISCUSSION

A working memory task was used to assess cognitive function and the underlying brain activation throughout the four phases of the migraine cycle across multiple spontaneous attacks in women with low-frequency episodic menstrual migraine without aura. We compared patients with a control group of healthy women while in the corresponding phases of their menstrual cycle. We found an overall preserved working memory performance in patients relative to controls across the cycle, paralleled by a similar activation of working memory related areas. Significant blunted BOLD response was identified in the hypothalamus of M patients during the postictal phase when compared to HC perimenstrual. The hypothalamus and the brainstem have been established as contributing drivers of migraine attacks and maintenance of the headache phase^5, 27, 31–34^, yet our results suggest that changes in the dynamics on at least one of these subcortical regions, namely the hypothalamus, could also be present after the attack.

The results obtained in the n-back task indicated no significant differences between groups nor phases in the behavioral measures of correct or incorrect response rate, and reaction times. Only a marginal trend when comparing incorrect responses between the perimenstrual and peri-ictal sessions was observed (*p* = 0.054), potentially influenced by the reduced sample size with more incorrect responses percent in postictal phase. This is in line with previous findings from two test-retest studies in migraine and control populations, comparing migraine during interictal and postictal phase through neuropsychological performance^28–29^, including the 2-back^28^. Both reported no significant impairment in cognitive performance during postictal phase nor related to controls. Yet, a study using an online version of the 2-back also observed a higher rate of incorrect responses in pain condition when compared to no pain^30^, which suggest that a bigger sample size might be necessary to infer more definite conclusions.

Brain activity observed during a working memory task in the M interictal sessions was comparable to what was observed in HC, and HC exhibited similar patterns of brain activation along the different stages of the menstrual cycle. These similarities applied both for the voxel-wise and ROI analyses involving cortical and subcortical brain areas. This is in line with what was described previously, although with another cognitive task, by Mathur and colleagues^7^.

We observed significant changes during the working memory task, with a main effect of subcortical brain activation in between migraine phases in the thalamus, brainstem, and hypothalamus. Specifically, this decrease in BOLD percent change was present in the hypothalamus during the postictal phase, when compared both to HC perimenstrual, M preictal and interictal phases.

Enhanced activity has been demonstrated in the brainstem^5^ and hypothalamus during the preictal phase^5, 27,31–35^, while brainstem-hypothalamic functional coupling has been described during preictal and ictal phases, both during resting state^35^ and nociceptive stimulation^27^. The latter was also observed during postictal phase, although not reaching statistical significance^27^. The only reported brain activity observed during the postictal phase was limited to the visual cortex, namely in response to pain stimulation^27^.

The observed decrease in subcortical BOLD activity in individuals experiencing cognitive demands during attacks may be attributed to compensatory processes. The abovementioned subcortical structures have been previously observed in the context of the execution of higher order cognitive functions. Under such demands, it would make sense for example to repress incoming signals from the thalamus, a pain encoding area, which otherwise has been described as active both for visual and verbal n-back tasks^36^. A study using auditory evoked potentials observed an interaction between the DLPC and the brainstem while performing an n-back task, where the higher the working memory load, the lower brainstem response was obtained^37^. They hypothesized that an inhibitory mechanism of the brainstem could prevent potential auditory distractions from the ongoing task^37^. This mechanism could potentially account for the observed correlation pattern between subcortical brain activity and performance. Specifically, as subcortical brain activity increases, there is a corresponding increase in the percentage of correct responses, a decrease in the percentage of incorrect responses, and a decrease in reaction times.

A significant reduction in BOLD response within the ACC was also observed during postictal phase when compared to the preictal phase. The anterior cingulate cortex plays a crucial role in cognitive and emotional processing due to its extensive connections, serving as a bridge between limbic and cortical regions, such as the orbito-prefrontal cortex^38^. Previous results obtained by our group in a different set of women with episodic migraine showed an increased BOLD activity in the orbito-prefrontal cortex, during the ictal phase when compared to the interictal phase, also linked to a verbal 2-back task^8^. A similar effect was also found in the present sample for the orbito-prefrontal cortex; however, it did not reach significance after correction for multiple comparisons. Taken together with the present findings, our results suggest that when performing a cognitive task during a migraine attack, cerebral areas related to pain processing and migraine pathophysiology may be interconnected in terms of function and have a mutual influence^39^ with cognitive top-down areas.

## Limitations

The primary constraint of this study were the methodological difficulties related to collecting data during spontaneous migraine attacks, which limited the available sample size. Due to our unique recruitment of female patients, it is not possible to generalize our findings to male subjects. Also, the confirmation of menstrual cycle stages was conducted by menstrual calendar, and not through hormone concentration techniques like saliva or urine. Both the migraine and control groups included participants who were taking contraceptive medication, potentially homogenizing the effects of the hormone cycle. Ultimately, six participants withdrew from the study at different stages of the protocol, resulting in their respective sessions being excluded from the statistical analysis for comparing in-between migraine phases.

## Conclusion

Brain activity in areas involved in cognitive top-down processes was linked to a working memory task across all phases of migraine and alike healthy participants. A postictal reduction in BOLD activity in the hypothalamus associated with the task may indicate a functional interplay between cognitive areas and the latter. This enhances our comprehension of the brain mechanisms involved in effectively managing cognitive demands during spontaneous migraine attacks, highlighting the significance of subcortical regions.

## Data Availability

All data produced in the present study are available upon reasonable request to the authors.

## Acknowledgements

We are thankful for the effort and commitment of the participants. We are also grateful to the administrative staff and neuroimaging technicians of Hospital da Luz for their assistance.

